# Association of Endovascular Thrombectomy Volume and Outcomes in Acute Ischemic Stroke: A National Inpatient Sample Study

**DOI:** 10.1101/2024.06.04.24308466

**Authors:** Lane Fry, Aaron Brake, Cody Heskett, Frank A. De Stefano, Ari Williams, Nashaat Majo, Catherine Lei, Abdul-Rahman Alkiswani, Kevin Le, Adam G. Rouse, Jeremy Peterson, Koji Ebersole

**Author notes:** **Corresponding Author:** Lane Fry, University of Kansas Medical Center, Department of Radiology, 3901 Rainbow Blvd, MS 4032, Kansas City, Kansas, 66160. Disclosures: None. Funding: None.

## Abstract

**Background:** Previous studies have reported a positive relationship between higher hospital endovascular thrombectomy (EVT) volume and shorter procedures, higher revascularization rates, and improved functional outcomes. We investigated the association between hospital EVT volume and clinical outcomes using the National Inpatient Sample (NIS) database from 2016-2020.

**Methods:** A cross-sectional analysis of the NIS examining the relationship of hospital EVT volume and outcomes was performed. All relevant clinical and demographic information was collected. The outcomes were favorable functional outcome (home without assistance), inpatient mortality, and intracerebral hemorrhage (ICH). Centers were classified as high-volume if they were in the top quintile of annual EVT volume. We performed univariate, multivariate, nearest neighbor matched analysis, and an exploratory annual case volume cutoff analysis.

**Results:** There were 114,640 patients who underwent EVT included in the sample. Of these, 24,415 (21.3%) were in the high-volume group. High-volume centers had higher rates of favorable functional outcome in univariate (OR 1.20, p < 0.001), multivariate (aOR 1.19, p = 0.003), and matched analysis (OR 1.14, p = 0.028). Prior to matching, lower rates of inpatient mortality (OR 0.83, p < 0.001). However, in univariate and matched analysis there were no differences between high and low-volume centers. There were no differences in ICH across all analyses. Functional benefit was first noted at ≥ 50 EVTs, but centers performing ≥ 175 EVTs had substantially higher functional benefit (aOR 1.42, p = 0.002).

**Conclusions:** Our analysis demonstrates increased hospital case volume is associated with a modest improvement in favorable functional outcomes in patients undergoing EVT for AIS. Attempts to identify procedural cut off values reveal likely improved functional outcomes beginning at 50 EVT per year, while this benefit seems to increase with increasing case volumes. These higher levels of case volumes do not lead to higher rates of inpatient mortality or ICH.

## Introduction

Endovascular thrombectomy (EVT) has emerged as the standard of care for patients presenting with acute ischemic stroke due to large vessel occlusion (LVO), demonstrating significant benefits in terms of revascularization, neurological recovery, and functional outcomes.^1,2^ The successful implementation of EVT relies on well-coordinated stroke systems of care, incorporating streamlined processes for patient selection, timely intervention, and optimal post-procedural management. Previous studies have reported a positive relationship between higher hospital EVT volume and shorter procedure times, higher revascularization rates, and improved functional outcomes.^3–8^ As EVT becomes increasingly integrated into routine clinical practice, understanding the impact of hospital case volume on patient outcomes is essential for optimizing stroke care.

Two prior administrative studies have examined the volume-outcome relationship for EVT in acute ischemic stroke.^3,4^ Each of these studies demonstrated benefit for patients at high-volume thrombectomy centers in terms of favorable functional outcome and inpatient mortality. However, the patient population in both studies were limited to only patients on Medicare and only encompassed data from 1-2 years.

In this study, we sought to investigate the association between hospital EVT volume and clinical outcomes using the National Inpatient Sample (NIS) database from 2016-2020. The NIS is a comprehensive and nationally representative dataset of inpatient hospital stays in the United States, including all payers. Our objective was to provide robust, population-level evidence to inform ongoing discussions regarding the optimization of EVT delivery and stroke care networks. We hypothesized that higher volume centers would have higher rates of favorable functional outcome when compared to lower volume centers.

## Methods

### Study Design

This is a cross-sectional retrospective analysis of United States hospital inpatient admissions examining the relationship of hospital EVT volume and outcomes. This study was conducted in accordance with STROBE guidelines.

### Data Source

The NIS is the largest national inpatient database in the United States, containing 7 million unweighted hospital admissions annually. It is developed and maintained by the Healthcare Cost and Utilization Project (HCUP) and consists of de-identified inpatient hospitalization data derived from billing and discharge information from the participating hospitals. Utilization of provided discharge weights reported by participating institutions allows for estimates of nationally representative statistics.

### Patient Selection

Weighted discharge NIS data from 2016-2020 was surveyed for patients admitted with primary *International Classification of Disease, Tenth Version*, (ICD-10) admission diagnosis of AIS (cerebral infarction - *ICD-10-CM* I63.xx). Endovascular thrombectomy (EVT) (*ICD-10-PR* 03CGxx, 03CHxx, 03CJxx, 03CKxx, 03CLxx, 03CPxx, 03CQxx, 037Gxx, 03Qxx) and intravenous tPA (IV-tPA) (*ICD-10-CM* Z92.82, *ICD-10-PR* 3E03316, 3E03317, 3E04316, 3E04317) procedural codes were then interrogated to determine treatment exposure. Patients were included if they were admitted for AIS and received EVT. An overview of patient selection is seen in **Supplemental Figure 1**.

**Figure 1.**
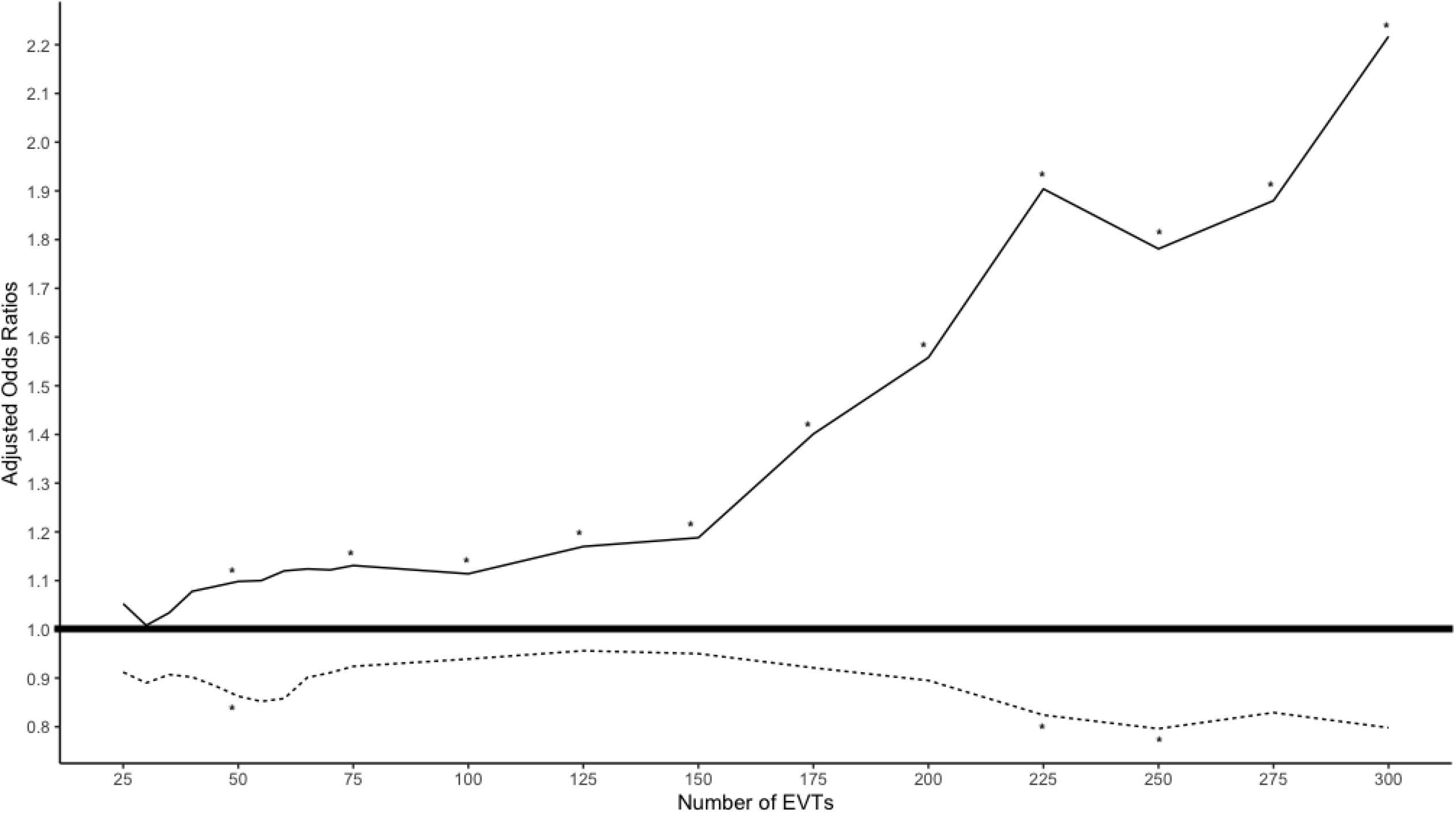
Line plots demonstrating the increase in total number of endovascular thrombectomies (EVT) performed and total centers performing EVT in the United States from 2016 to 2020.

### Demographic and Clinical Characteristics

Baseline demographic data extracted from the NIS database was collected including age, sex, race, primary payer status, hospital location, and teaching status. Comorbidity burden was assessed by utilizing the Charlson comorbidity index.^9,10^ We identified markers of stroke severity based on previous NIS studies analyzing EVT outcomes.^11,12^ Markers of stroke severity included ICD-10 diagnoses related to level of consciousness (stupor and coma), infarct size (cerebral edema and herniation), neurological deficits (neglect, dysphagia, and aphasia), and reliance upon respiratory or nutritional support (mechanical ventilation, nasogastric tube, and parenteral nutrition). Additionally, the proportion of total AIS admissions undergoing EVT were collected.

### Outcome Measures

The primary clinical end point of this study was favorable functional outcome, defined as a routine discharge to home without assistance. This discharge disposition has been demonstrated to have a strong correlation with mRS ≤ 2 at 90 days.^12–14^ Secondary outcomes were the rates of ICH and inpatient mortality between the treatment groups. Additionally, we investigated all discharge destinations (Short-Term Rehab Hospital, Skilled Nursing/Intermediate Care Facility, Home Health Care) available in the NIS for each group. The length of stay (LOS) and the total charges from the admission were also analyzed.

### Determination of EVT Volume Status and Hospital Characteristics

Utilizing the NIS HOSP_NIS variable we counted the number of EVT procedures performed at each unique hospital for each year included in the analysis. High-volume centers were defined as those being in the top 20^th^ percentile of EVT volume. This volume determination has been used prior in other national database analysis.^15,16^

To further explore potential volume cut-off values where outcomes were significantly improved, we used similar methodology as Stein et al.^4^ We compared favorable functional outcomes and inpatient mortality measures between increasing intervals of EVT procedures performed at a hospital. We started at a volume of 25 EVTs and increased by 25 EVTs until we reached a threshold of 300 EVTs performed. Once we discovered statistical differences between the high and low-volume groups, we utilized increments of 5 EVTs to determine a more precise cutoff point.

### Propensity Score Matching

Patients were matched 1:1 between high and low-volume centers utilizing nearest neighbor matching using logit distance. Patients were matched based on demographic (age, sex, minority status, and location/teaching status of the hospital), comorbidity burden (Charlson Comorbidity Index, Hypertension, Intracranial Atherosclerotic Disease (ICAD), Atrial Fibrillation, and Diabetes), stroke severity (as described above), and IV-tPA status.

### Statistical Analysis

Statistical analyses were performed accounting for the sampling design of the NIS, with appropriate strata, weights, and clusters according to HCUP guidelines. Statistical analyses were conducted with RStudio (Posit Software, Boston, MA, USA). Propensity matching was performed with the “MatchIt” package.^17^ All complex survey samples, including regression analysis and statistical tests, were performed with the “Survey” package.^18^ Survey weighted t-tests and chi-squared analysis were performed to compare means and association of categorical variables, respectively. Univariate and multivariate models were used to compare the high-volume group to the low-volume group with all outcome measures and discharge dispositions. Variables with a significance level < 0.1 were included in multivariate analysis. This was done with an unmatched cohort and then again with the matched cohort model. We assessed the trends of the number of EVTs performed nationwide and the number of centers offering EVT during the study period utilizing a Pearson’s correlation coefficient. We assumed the number of centers was distributed as a Poisson random variable, and thus performed a square root transformation to approximate a normal distribution before calculating the correlation. An alpha threshold of 0.05 was selected to define significance, with all p-values being 2-sided.

## Results

Centers grouped in the top quintile of EVT volume performed ≥ 115 EVTs in a given year. Centers performing < 115 EVTs in a given year were analyzed as the low-volume cohort. After aggregating the number of centers in individual years, there were 158 high-volume centers and 2,580 low-volume centers during the study period. The total number of centers performing EVT increased over time (*r* = 0.960, p =0.009). The numbers of high-volume (*r* = 0.961, p =0.009) and low-volume centers (*r* = 0.952, p =0.013) both increased during the study period. The number of EVTs performed increased at both high (*r* = 0.963, p =0.008) and low-volume centers (*r* = 0.988, p =0.002). The trend of centers and number of EVTs performed is seen in **Figure 1**.

Before propensity score matching, there were 114,640 patients who underwent EVT included in the sample. Of these, 24,415 (21.3%) were in the high-volume group. An overview of patient groups is seen in **Supplemental Figure 1**. The mean (± SD) number of EVTs performed at high-volume centers was 166 (± 49), while there was an average of 55 (± 28) EVTs performed at low-volume centers. There were higher rates of IV-tPA at low-volume centers (36.73% vs. 32.77%, p < 0.001). High-volume centers had lower rates of atrial fibrillation (41.80% vs. 44.95%, p < 0.001) and younger patients (68.66 vs. 69.34, p = 0.022). These findings are summarized in **Table 1**.

**Table 1.**
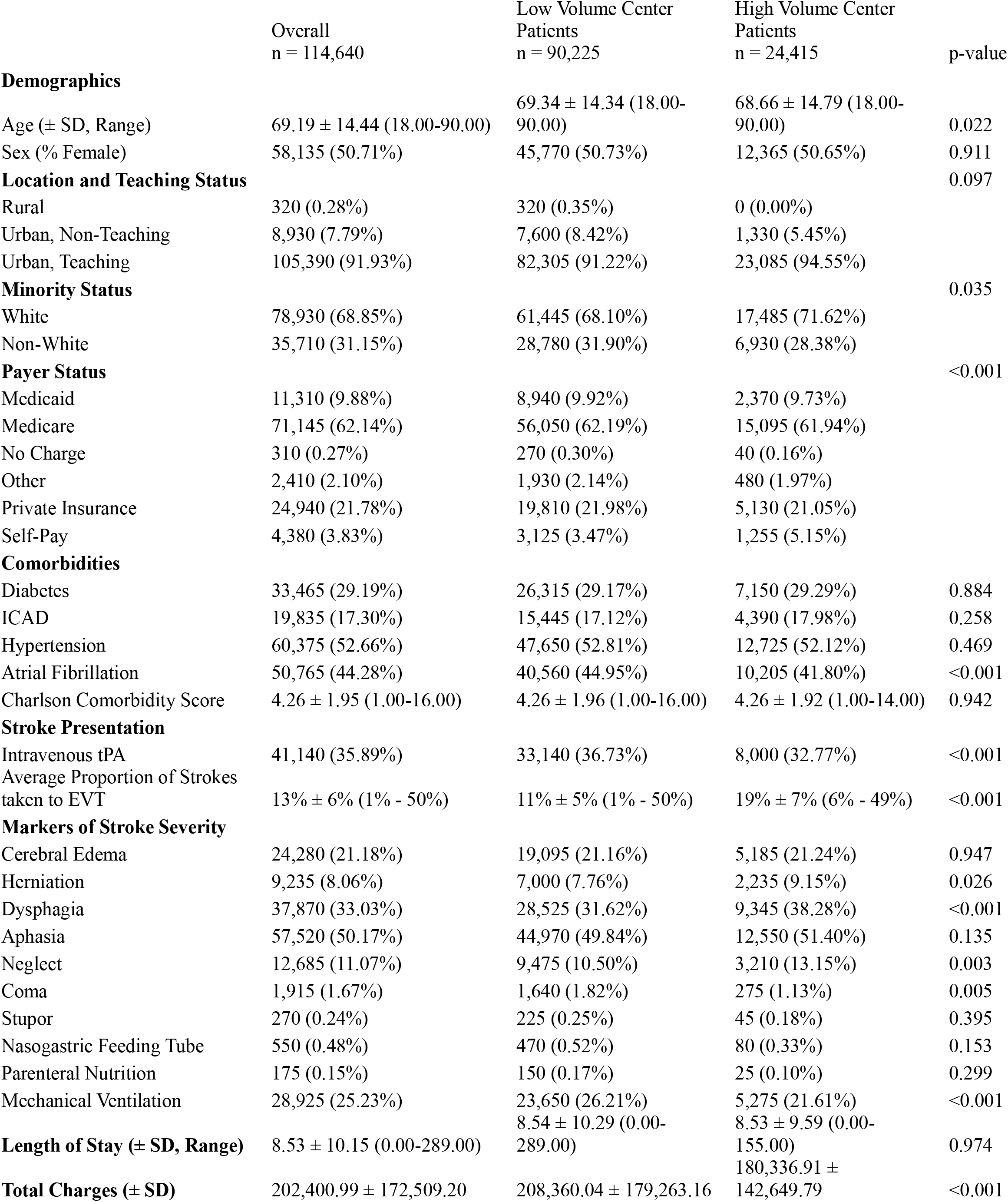
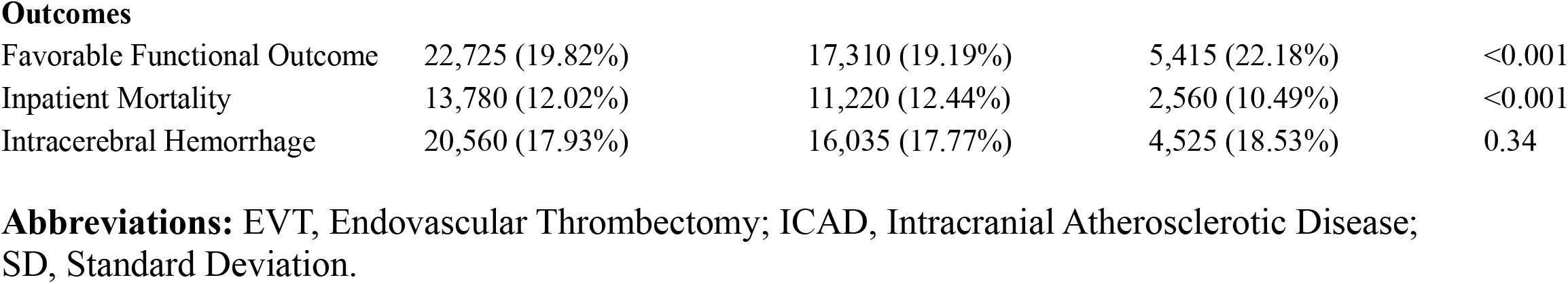
Demographic, clinical, and outcome measures of unmatched cohort.

After nearest neighbor matching, there were 24,415 patients in each group. There were no statistical differences in age, sex, comorbid conditions, utilization of tPA, or markers of stroke severity in the matched groups. The nearest neighbor matching adequately matched comorbidities, markers of stroke severity, and utilization of IV-tPA. High-volume centers performed EVT in a higher proportion of stroke patients than low-volume centers (19% vs 12%, p < 0.001). The results after matching are summarized in **Table 2**.

**Table 2.** Demographic, clinical, and outcome measures after nearest neighbor matching.

|  | Overall<br>n = 48,830 | Low Volume Center<br>Patients<br>n = 24,415 | High Volume Center<br>Patients<br>n = 24,415 | p-value |
| --- | --- | --- | --- | --- |
| **Demographics** |  |  |  |  |
| Age (± SD, Range) | 68.84 ± 14.68 (18.00-90.00) | 69.02 ± 14.57 (18.00-90.00) | 68.66 ± 14.79 (18.00-90.00) | 0.295 |
| Sex (% Female) | 25,085 (51.37%) | 12,720 (52.10%) | 12,365 (50.65%) | 0.129 |
| **Location and Teaching Status** |  |  |  |  |
| Rural | 0 (0.00%) | 0 (0.00%) | 0 (0.00%) | ><br>0.999 |
| Urban, Non-Teaching | 2,635 (5.40%) | 1,305 (5.35%) | 1,330 (5.45%) |  |
| Urban, Teaching | 46,195 (94.60%) | 23,110 (94.65%) | 23,085 (94.55%) |  |
| **Minority Status** |  |  |  |  |
| White | 35,245 (72.18%) | 17,760 (72.74%) | 17,485 (71.62%) | 0.505 |
| Non-White | 13,585 (27.82%) | 6,655 (27.26%) | 6,930 (28.38%) |  |
| **Comorbidities** |  |  |  |  |
| Diabetes | 14,165 (29.01%) | 7,015 (28.73%) | 7,150 (29.29%) | 0.58 |
| ICAD | 8,495 (17.40%) | 4,105 (16.81%) | 4,390 (17.98%) | 0.191 |
| Hypertension | 25,380 (51.98%) | 12,655 (51.83%) | 12,725 (52.12%) | 0.8 |
| Atrial Fibrillation | 20,760 (42.51%) | 10,555 (43.23%) | 10,205 (41.80%) | 0.182 |
| Charlson Comorbidity Score (± SD, Range) | 4.26 ± 1.95 (1.00-15.00) | 4.25 ± 1.98 (1.00-15.00) | 4.26 ± 1.92 (1.00-14.00) | 0.864 |
| **Stroke Presentation** |  |  |  |  |
| Intravenous tPA | 16,190 (33.16%) | 8,190 (33.54%) | 8,000 (32.77%) | 0.52 |
| Average Proportion of Strokes taken to EVT | 15% ± 7% (1% - 49%) | 12% ± 5% (1% - 33%) | 19% ± 7% (6% - 49%) | <0.001 |
| **Markers of Stroke Severity** |  |  |  |  |
| Cerebral Edema | 10,075 (20.63%) | 4,890 (20.03%) | 5,185 (21.24%) | 0.312 |
| Herniation | 4,410 (9.03%) | 2,175 (8.91%) | 2,235 (9.15%) | 0.741 |
| Dysphagia | 18,420 (37.72%) | 9,075 (37.17%) | 9,345 (38.28%) | 0.467 |
| Aphasia | 25,085 (51.37%) | 12,535 (51.34%) | 12,550 (51.40%) | 0.959 |
| Neglect | 6,560 (13.43%) | 3,350 (13.72%) | 3,210 (13.15%) | 0.593 |
| Coma | 540 (1.11%) | 265 (1.09%) | 275 (1.13%) | 0.858 |
| Stupor | 75 (0.15%) | 30 (0.12%) | 45 (0.18%) | 0.43 |
| Nasogastric Feeding Tube | 145 (0.30%) | 65 (0.27%) | 80 (0.33%) | 0.617 |
| Parenteral Nutrition | 50 (0.10%) | 25 (0.10%) | 25 (0.10%) | >0.999 |
| Mechanical Ventilation | 10,220 (20.93%) | 4,945 (20.25%) | 5,275 (21.61%) | 0.265 |
| **Length of Stay (± SD, Range)** | 8.36 ± 9.11 (0.00-155.00) | 8.20 ± 8.61 (0.00-147.00) | 8.53 ± 9.59 (0.00-155.00) | 0.153 |
| **Total Charges (± SD)** | 189,654.70 ± 153,334.78 | 198,974.41 ± 162,806.47 | 180,336.91 ± 142,649.79 | 0.013 |
| **Outcomes** |  |  |  |  |
| Favorable Functional Outcome | 10,290 (21.07%) | 4,875 (19.97%) | 5,415 (22.18%) | 0.028 |
| Inpatient Mortality | 5,180 (10.61%) | 2,620 (10.73%) | 2,560 (10.49%) | 0.709 |
| Intracerebral Hemorrhage | 8,810 (18.04%) | 4,285 (17.55%) | 4,525 (18.53%) | 0.287 |

**Abbreviations:** EVT, Endovascular Thrombectomy; ICAD, Intracranial Atherosclerotic Disease; SD, Standard Deviation.

Prior to matching, the high-volume centers had higher rates of favorable functional outcome, as defined by discharge to home without assistance (22.18% vs. 19.19%; OR 1.20, 95% CI [1.08-1.33]; p < 0.001) and lower rates of inpatient mortality (10.49% vs. 12.44%; OR 0.83, 95% CI [0.74-0.92]; p < 0.001), but there was no difference in rates of ICH (18.53% vs. 17.77%; OR 1.05, 95% CI [0.95-1.17]; p = 0.340). After matching, univariate analysis revealed high-volume centers outperformed low-volume centers in terms of favorable functional outcome (22.18% vs. 19.97%; OR 1.14, 95% CI [1.01-1.29]; p = 0.028). There was no difference in inpatient mortality (10.49% vs. 10.73%; OR 0.97, 95% CI [0.85 - 1.12]; p = 0.709) or rates of ICH (18.53% vs. 17.55%; OR 1.07, 95% CI [0.95-1.21]; p = 0.287) after matching. Multivariate analysis of unmatched data demonstrated a persistent benefit for favorable functional outcome for the high-volume group (aOR 1.19 95% CI [1.06 - 1.34], p = 0.003) with no differences in inpatient mortality and ICH. Examining discharge dispositions, there were no differences in all other discharge destinations (Rehab, Skilled Nursing Facility, or Home with assistance) in univariate or multivariate models. There were no differences in LOS before and after matching. However, the mean total charges for patients at high-volume centers was significantly less in both the unmatched ($180,336.91 vs. $208,360, p < 0.001) and matched ($180,336.91 vs. $198,974.41, p = 0.013) cohorts. All outcome models are summarized in **Table 3**.

**Table 3.** Univariate, Multivariate, and Matched Logistic Regression Models of Outcome Measures and Discharge Dispositions comparing High Volume Centers to Low Volume Centers.

|  | Low Volume Center Patients | High Volume Center Patients | Odds Ratios (95% Confidence Interval) |  |  |  |  |  |
| --- | --- | --- | --- | --- | --- | --- | --- | --- |
|  |  |  | Unmatched Univariate | p-value | Matched Univariate | p-value | Unmatched Multivariate* | p-value |
| Favorable Functional Outcome (High vs. Low Volume) | 17,310 (19.19%) | 5,415 (22.18%) | 1.20 (1.06 - 1.33) | < 0.001 | 1.14 (1.01 - 1.29) | 0.028 | 1.19 (1.06 - 1.34) | 0.003 |
| Inpatient Mortality (High vs. Low Volume) | 11,220 (12.44%) | 2,560 (10.49%) | 0.83 (0.74 - 0.92) | < 0.001 | 0.97 (0.85 - 1.12) | 0.709 | 0.92 (0.81 - 1.04) | 0.184 |
| Intracerebral Hemorrhage (High vs. Low Volume) | 16,035 (17.77%) | 4,525 (18.53%) | 1.05 (0.95 - 1.17) | 0.34 | 1.07 (0.95 - 1.21) | 0.287 | 1.04 (0.93 - 1.15) | 0.574 |
| Short-term Rehab Hospital | 3,020 (3.35%) | 660 (2.70%) | 0.80 (0.46 - 1.40) | 0.439 | 0.93 (0.52 - 1.65) | 0.798 | 0.87 (0.50 - 1.52) | 0.629 |
| Skilled Nursing Facility | 49,630 (55.01%) | 13,180 (53.98%) | 0.96 (0.88 - 1.04) | 0.328 | 0.92 (0.83 - 1.01) | 0.08 | 0.92 (0.84 - 1.00) | 0.053 |
| Home Health | 9,045 (10.02%) | 2,600 (10.65%) | 1.07 (0.95 - 1.21) | 0.268 | 1.04 (0.90 - 1.20) | 0.59 | 1.08 (0.96 - 1.22) | 0.205 |

\*Adjusted for Age, Atrial Fibrillation, Intravenous tPA, Location and Teaching Status, Minority Status, Herniation, Dysphagia, Neglect, Coma, Mechanical Ventilation, and Proportion of Strokes treated with EVT at a specific Hospital

In our exploratory volume cut-off analysis, a difference in the combined metric of favorable functional outcome and inpatient mortality manifested between 25 and 50 EVTs. A mortality benefit alone manifested at 45 EVTs while higher odds of favorable functional outcome alone manifested at 50 EVTs performed annually. The volume cut-off analysis is seen in **Figure 2**.

**Figure 2.**
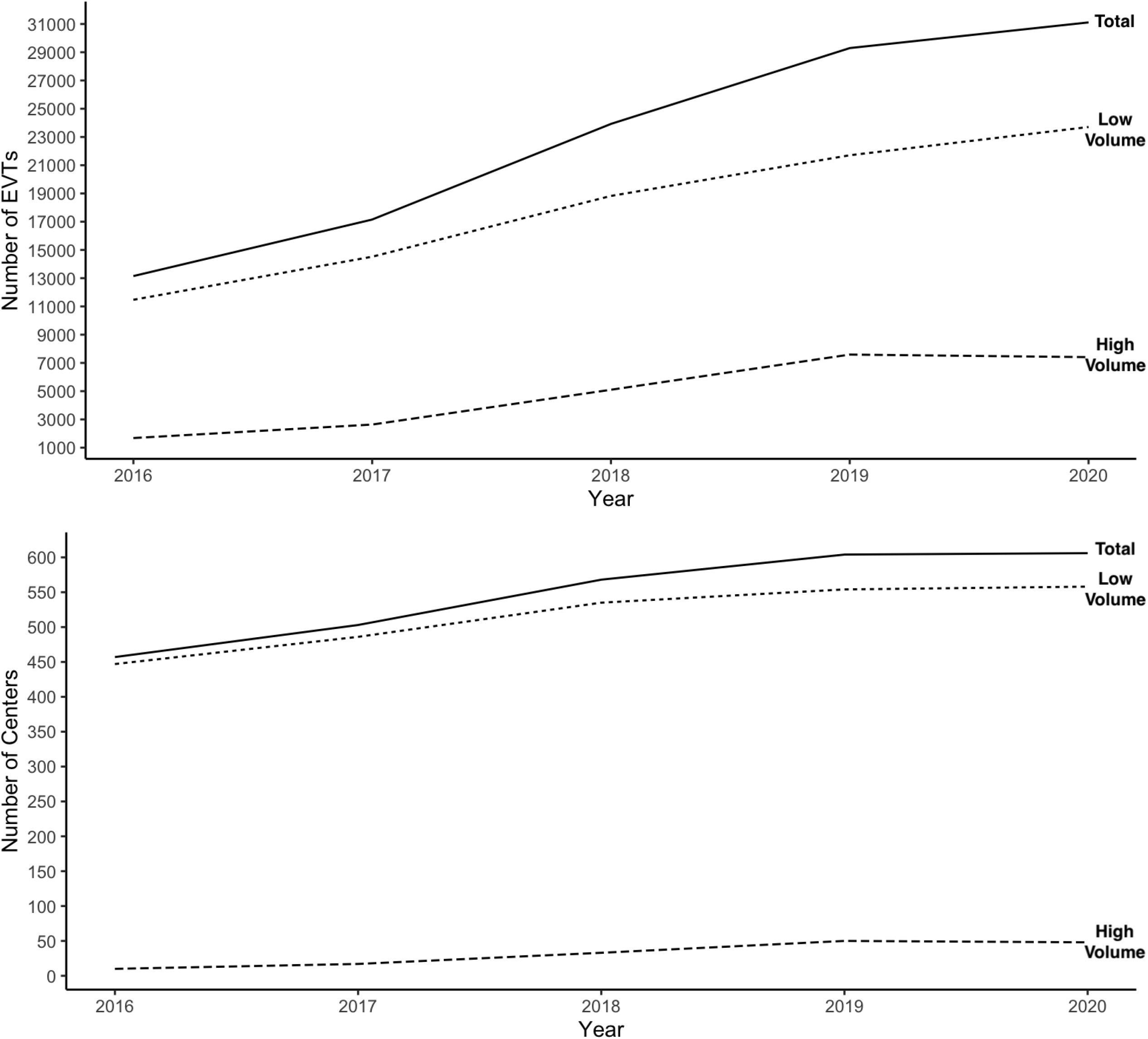
Line plots demonstrating between dichotomized volume thresholds and outcomes in multivariate models for hospital case volume. Models adjusted age, atrial fibrillation, intravenous tPA, location and teaching status, minority status, herniation, dysphagia, neglect, coma, mechanical ventilation, and proportion of strokes treated with endovascular thrombectomy at a specific hospital. * Denotes a statistically difference in adjusted odds ratios

There was a notable increase in the odds of favorable functional outcome in patients treated at centers performing ≥ 175 EVTs per year, identifying a group we labeled “super high-volume centers.” This finding prompted an additional multivariate sub-analysis comparing three groups, low volume vs. high-volume vs. super high-volume centers. Compared to low-volume centers, super-high-volume centers had higher odds of favorable functional outcomes (aOR 1.42, 95% CI [1.13 - 1.78], p = 0.002) while high-volume centers did not differ from the low-volume centers (aOR 1.09, 95% CI [0.97 - 1.22], p = 0.158). Neither the super-high (aOR 0.91, 95% CI [0.75 - 1.11], p = 0.341) nor the high-volume (aOR 0.92, 95% CI [0.79 - 1.07], p = 0.291) centers outperformed low-volume centers in terms of mortality. There was also no difference in rates of ICH between super-high (aOR 1.14, 95% CI [0.93-1.38], p = 0.209) nor high-volume (aOR 0.99, 95% CI [0.88-1.11], p=0.801) centers compared to low-volume centers. These findings are summarized in **Table 4**.

**Table 4.**
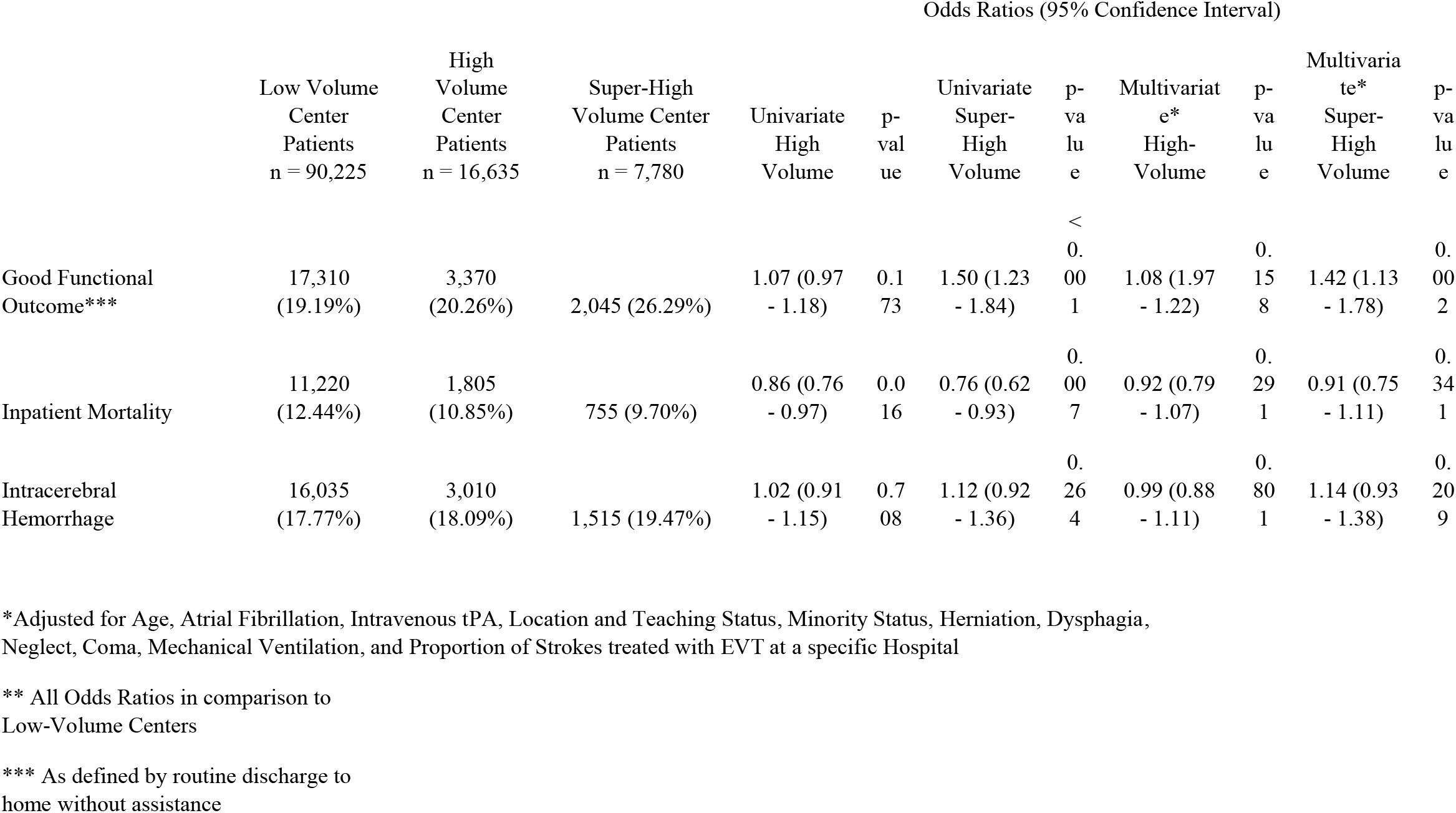
Univariate and Multivariate Logistic Regression Models comparing Super-High Volume vs. High Volume vs. Low Volume Centers.

## Discussion

An association between case volume and improved patient outcomes is well documented in the literature for percutaneous coronary intervention for acute coronary syndromes.^19^ A similar finding has been demonstrated among centers performing mechanical thrombectomy for acute stroke.^3,4^ For example, Stein et al., using Medicare/Medicaid data, estimated the annual hospital volume for outcome benefit and mortality benefit were 26.2 and 44.8, respectively.^4^ In another analysis of CMS data, Waqas et al. demonstrated that hospital EVT volume was associated with lower mortality and higher discharge to home rates, identifying the two central quartiles of mortality and discharge to home rates to be 24 and 23 respectively.^3^ Accordingly, neuroendovascular certification and training standards reflect the intuitive understanding that a relationship between case volume and outcomes likely exists. For example, as of 2021, The Joint Commission recommends 15 thrombectomies per year or 30 over 2 years to meet criteria for Thrombectomy-capable Stroke Center (TSC) or a Comprehensive Stroke Center (CSC) certification. The Society of Neurological Surgeons (SNS) and its Committee for Advanced Subspecialty Training (CAST) has defined a minimum number of cases performed for competency in neuroendovascular surgery including at least 30 acute ischemic stroke interventions.^20^

Consistent with prior studies and certification standards, our analysis of the NIS database also finds that higher hospital thrombectomy case volume is positively associated with higher rates of favorable functional outcome. Unlike prior studies, our analysis includes all patients who received EVT from 2016-2020, as reported in the database, regardless of payor source, yielding a larger sample size than previous studies and one that more closely reflects real-world populations. In our analysis, favorable functional outcome was defined as routine discharge to home without assistance, which strongly correlates to an mRS ≤ 2.^12–14^ Thus the present study analyzes patients achieving the very best outcomes after intervention. The positive association between case volume and favorable functional outcome was persistent across all analyses. The benefit first manifests at ≥ 50 EVTs and demonstrates increasing benefit with higher annual case volume. In a sub-analysis however, this effect appears driven by the outcomes achieved by super-high-volume centers, those performing ≥ 175 EVTs in a year. Centers performing ≥ 175 EVTs annually outperformed lower volume centers in achieving very favorable outcomes by 6-7%. Further, the super-high volume high-volume centers referred a higher proportion of patients to mechanical thrombectomy, exposing more candidates to the benefit. The functional benefit is persistent after successful matching of patient groups and utilizing multivariate logistic regression. While a broader definition of “benefit” would undoubtedly yield a lower case volume requirement, perhaps even substantially so, our analysis suggests that the likelihood of achieving a very favorable outcome after mechanical thrombectomy is best at the very highest-volume centers.

Consistent with prior studies, our analysis finds that mechanical thrombectomy is safe. In unmatched, univariate analysis, higher volume centers had lower rates of inpatient mortality, but this difference was not observed after matching nor in adjusted analyses. In matched univariate analysis, super-high-volume centers demonstrated a mortality benefit compared to low-volume centers, but the difference was not observed in matched multivariate analysis. Meanwhile, there was no identifiable association between rates of ICH and thrombectomy volume status in any analysis. In other words, our analysis demonstrates that low volume centers achieved the same safety outcomes as even the super-high-volume centers. Viewed another way, our study finds that the super-high-volume centers matched the safety outcomes of lower-volume centers, despite a higher proportion of stroke patients being referred from thrombectomy, and more patients treated outside the tPA window.

Lastly, our analysis identified an apparent cost savings for patients treated at higher volume centers. Existing data reporting EVT procedure volume and incurred cost per patient does not exist to this point. Previous publications have only examined patient outcomes as a function of hospital volume.^3–5^ Our matched analysis demonstrates a nearly $18,000 savings per patient, on average, when treated at a high-volume center. Similar findings have been seen in other procedural disciplines with high-volume centers more likely to have lower costs and improved outcomes.^21,22^

## Limitations

This study is primarily limited by the inherent nature of sampling from a large national database and retrospective study design. The NIS collects a sample of discharged patient data from each hospital partaking in the HCUP consortium. As a sample driven database, the NIS is not able to give exact numbers of cases per institution. This means that our value estimation for cutoff data is based on hospital sample data and is likely under-estimating the true number of cases performed by each hospital. This has significant implications in terms of numeric derivatives (such as cutoff values for outcomes). However, as a sample database we are still able to reliably group hospitals into quintiles based on relative case volume. The NIS is an administrative database, reliant on coding via the ICD-10 system. Patients that may not represent our intended sample population may inadvertently be included due to coding errors. While it is less likely that coding mistakes may occur for inpatient mortality and disposition status, our sample is not immune from potential errors. Additionally, the HOSP_NIS variable does not carry over from year-to-year, so we are unable to assess if low-volume centers are increasing their volumes over time. Further, differing stroke triage systems exist and may alter the outcomes for stroke patients, but our study is not equipped to examine the effects of these systems as it relates to patient outcomes.^23^

## Conclusion

Our analysis demonstrates increased hospital case volume is associated with a modest improvement in favorable functional outcomes in patients undergoing EVT for AIS. Attempts to identify procedural cut off values reveal likely improved functional outcomes beginning at 50 EVT per year, while this benefit seems to increase with increasing case volumes. These higher levels of case volumes do not lead to higher rates of inpatient mortality or ICH. Higher volume centers were also associated with decreased costs per patient. These findings further validate the relationship between procedural volume and outcomes and should assist in construction of policies of optimal stroke-care management and credentialing of potential comprehensive stroke centers.

## Data Availability

Data available to the public through the Healthcare Utilization Project.

